# Has COVID-19 Hurt Resident Education? A network-wide resident survey on education and experience during the pandemic

**DOI:** 10.1101/2020.08.13.20171256

**Authors:** Alexander Ostapenko, Samantha McPeck, Shawn Liechty, Daniel Kleiner

**Author notes:** Correspondences should be addressed to: Daniel Kleiner, MD, Department of General Surgery, Danbury Hospital. 24 Hospital Ave, Danbury CT 06810.

## Abstract

**Purpose:** As the COVID-19 pandemic continues to evolve, the healthcare system has been forced to adapt in myriad ways. Residents have faced significant changes in work schedules, deployment to COVID-19 units, and alterations to didactics. This study aims to identify the effects of the COVID-19 pandemic on resident perception of their own education within the Nuvance Health Network.

**Methods:** We conducted an observational study assessing resident perception of changes in education and lifestyle during the COVID-19 pandemic. A survey was developed to assess the quality and quantity of resident education during this time and administered anonymously to all residents within the healthcare network.

**Results:** Eighty-four (68%) residents responded to the survey from five different specialties, including general surgery, internal medicine, obstetrics and gynecology, pathology, and radiology. The average change in hours per week performing clinical work was −5.6 hours (SD=16.8), in time studying was +0.02 hours (SD=4.6), in weekly didactics was −1.7 hours (SD=3.1), and in attending involvement was −1.2 hours (SD=2.3). Additionally, 32% of residents expressed concern that the pandemic has diminished their preparedness to become an attending, 13% expressed concern about completing graduation requirements, and 3% felt they would need an additional year of training.

**Conclusions:** During the COVID-19 pandemic thus far, residents perceived that time spent on organized didactics/conferences decreased and that attending physicians are less involved in education. Furthermore, the majority of residents felt that the quality of didactic education diminished as a result of the pandemic. Surprisingly, while many residents expressed concerns about being prepared to become an attending, few were concerned about completing graduation requirements or needing an extra year of education. In light of these findings, it is critical to devote attention to the effects of the pandemic on residents’ professional trajectories and create innovative opportunities for improving education during this challenging time.

## Introduction

The COVID-19 pandemic has led to business closures, shelter in place restrictions, and mandated social distancing throughout the country. Nuvance Health network is a consortium of hospitals in the Hudson Valley region of New York and the western Connecticut region, an area considered to be one of the epicenters of the first wave of the pandemic from March-May of 2020. Within the healthcare system, changes were rapidly made to the allocation of resources, workflows, and daily operations, from the cancellation of non-emergent operations to the transition of many sectors to Telehealth medicine^1,2,3^. In the short term, the effects of these swift adaptations on residents and fellows have been innumerable, including alterations in work schedules and duty hour limitations, deployment to units caring for patients with COVID-19, and significant modifications in resident education. Many institutions have transitioned to remote academic sessions, local and national research conferences have been postponed or cancelled, and requirements for standardized exams have changed^4^. The response to current events has varied across states, healthcare systems, and hospitals. We aim to determine the effects of the pandemic on resident education and experience at our healthcare network, hypothesizing that within our network effects will vary across specialties based on program-specific changes to the prior status quo.

## Materials and Methods

We developed a survey with the aim of assessing changes in resident education, experience, and lifestyle. This survey was administered anonymously through SurveyMonkey® to all residents within the Nuvance Health network. This included obstetrics and gynecology, pathology, internal medicine, general surgery, and radiology residencies. The survey was open for a three-week period from June 9^th^ through June 26^th^, 2020. For questions comparing amount of time spent on clinical duties, studying individually, weekly didactics, and attending involvement before and during the pandemic, we calculated the perceived difference, delta, in hours for each individual response. Mean and standard deviation for deltas were calculated for each question. A portion of the survey was in the form of five-point Leikert questions. The responses for those questions were separated by specialty, and the percentage of residents responding positively as “agree” or “strongly agree” was calculated. Our Institutional Review Board deemed this study exempt (IRB# 2012208337) and waved the need for participant consent.

## Results

A total of eighty-four residents completed the survey (Supplemental Table 1). The response rate per specialty was 60% for internal medicine, 91% for general surgery, 67% for obstetrics and gynecology, 75% for pathology, and 50% for radiology. Of the respondents: 43 (52%) were internal medicine, 20 (24%) general surgery, 10 (11%) obstetrics and gynecology, 6 (7%) pathology, and 6 (7%) radiology residents respectively. Including all residents in the 2019-2020 academic year within the Nuvance Health Network, the response rate to the survey was 68.3%. However, the questionnaire was administered at the time of graduation of senior residents, therefore it is likely that many of these residents accounted for missing responses. Excluding all seniors from possible responses results in a response rate of 96.6%; therefore, we estimate that the true response rate was between 68.3% and 96.6%.

On average, residents at Nuvance Health reported spending 29.3-39.3 hours per week carrying for COVID-19 patients, with internal medicine residents reporting the most hours between 43.3-53.3 (Supplemental Table 2). On average, residents perceived spending 5.6 fewer hours (SD=16.8hours) per week performing clinical work during the pandemic (Table 1). However, when analyzed by specialty, the difference in clinical work was quite variable. Internal medicine residents reported an increase of 5.1 hours per week, while general surgery and obstetrics and gynecology reported a decrease of 17 hours and 24.4 hours per week respectively. Similarly, the average difference in amount of time studying independently was grossly unchanged before and during the pandemic (mean difference= 0.02 hours; SD= 4.6 hours) (Table 2). However, there were significant differences when examined by specialty, with only internal medicine residents reporting less time spent studying.

**Table 1:** Average hours per week spent performing clinical work before and during the <u>pandemic by specialty</u>. Standard deviation is listed in parenthesis.

| Specialty | Before pandemic | During pandemic | Average Difference |
| --- | --- | --- | --- |
| Internal medicine | 64.7(15.2) | 69(16.0) | + 5.1(9.3) |
| General Surgery | 80.5(14) | 63.5(16.0) | - 17(16.2) |
| Obstetrics and gynecology | 78.8(6.0) | 54.4(8.8) | - 24.4(10.1) |
| Pathology | 41.6(25.6) | 35(28.1) | - 6.7(16.3) |
| Radiology | 58.3(9.8) | 43.3(13.6) | -15.0(16.4) |
| Overall | 67.9(18.1) | 62.2(19.2) | -5.6(16.8) |

**Table 2:** Average hours per week spent <u>studying individually</u> before and during the pandemic by specialty. Standard deviation is listed in parenthesis.

| Specialty | Before pandemic | During pandemic | Average Difference |
| --- | --- | --- | --- |
| Internal medicine | 5.9(3.1) | 3.1(2.3) | - 2.8(2.8) |
| General Surgery | 6.1(3.1) | 7.7(4.7) | + 1.6(4.7) |
| Obstetrics and gynecology | 4.44(1.9) | 10.2(4.0) | + 5.8(2.9) |
| Pathology | 3(1.7) | 5.7(5.3) | +2.7(3.9) |
| Radiology | 8.3(4.3) | 12(4.5) | +3.7(3.4) |
| Overall | 5.8(3.2) | 5.8(4.6) | +0.02(4.6) |

Fewer scheduled academic weekly conferences were reported, with an average decrease of 1.7 hours per week during COVID-19 (SD=3.1), (Table 3). Across specialties, residents also perceived less involvement from attending physicians during the pandemic, with a perceived decrease of 1.2 hours per week (SD=2.3) (Supplemental Table 3). Overall, there was no strong consensus regarding changes in the quality of didactics: 53.5% reported it decreased and 23.8% reported it improved (Table 4). However, when responses were analyzed by specialty, the responses diverged. No internal medicine residents reported an improvement in quality (80% perceived a decrease in quality of their lectures), while 100% of pathology residents and 55% of obstetrics and gynecology residents reported improvement.

**Table 3:**
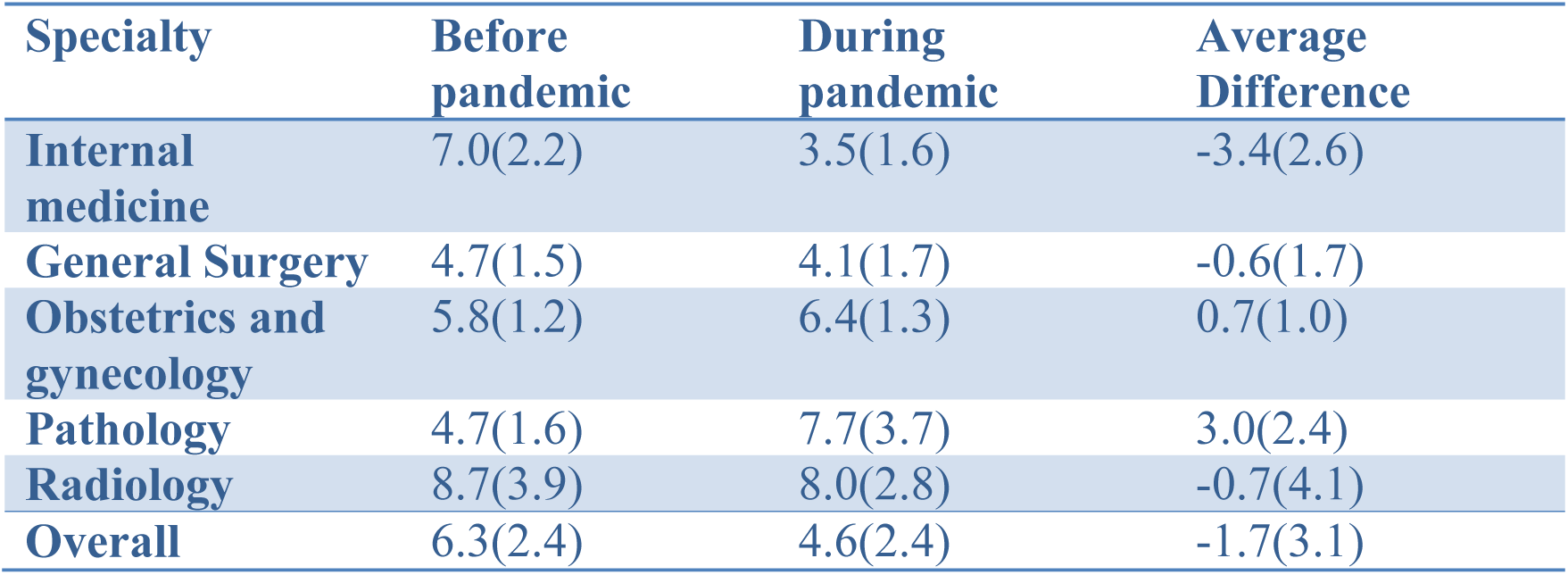
Average hours per week spent on <u>didactics and academic conferences</u> before and during the pandemic by specialty. Standard deviation is listed in parenthesis.

**Table 4:** Number and percentage of responses to “The quality of didactics increased during the pandemic.” Responders that chose “disagree” or “strongly disagree” perceived a decrease in quality, while those who chose “agree” or “strongly agree” perceived an improvement. Responses are broken down by specialty.

| Specialty | Decrease in quality | Percentage of total within specialty | Increase in quality | Percentage of total within specialty |
| --- | --- | --- | --- | --- |
| **Internal medicine** | 36 | 83.7% | 0 | 0.0% |
| **General Surgery** | 7 | 35.0% | 7 | 35.0% |
| **Obstetrics and gynecology** | 1 | 11.0% | 5 | 55.0% |
| **Pathology** | 0 | 0.0% | 6 | 100.0% |
| **Radiology** | 1 | 16.7% | 2 | 33.3% |
| **Overall** | 45 | 53.6% | 20 | 23.8% |

Regarding the effect of the changes in academics on overall education, 32.1% of residents expressed concern that the pandemic has diminished their preparedness to become an attending (Table 5). Despite this finding, few expressed concern about completing graduation requirements (13.0%) or felt they would need an additional year of training (3%).

**Table 5:** Preparedness to become an attending. Number and percentage of responses who chose “Agree” or “Strongly agree” to the respective questions.

| Question | Number of residents | Percentage |
| --- | --- | --- |
| Concerned about preparedness to become an attending | 27 | 32.1% |
| Concerned about completing graduation requirements | 11 | 13.0% |
| Concerned about requiring an extra year of training | 3 | 3.6% |

## Discussion

COVID-19 has affected the United States healthcare system in profound and unanticipated ways, and it will likely take years to understand and quantify the full scope of its effects. Thus far, many publications have emerged with observations, recommendations, and trials regarding patient care, specific treatment modalities, and occupational safety. As of yet, little literature is available the pandemic’s effects on resident education^5^. Residency programs across the country have been forced to make sweeping changes to their customary operations, and have appropriately maintained focus on safety and patient care.

One side effect of many of these changes has been a decrease in protected time for academics.^6^ Overall, at our institution there was a 1.7-hour per week decrease in the amount of time spent on academic conferences and didactics (Table 3). Only pathology and obstetrics and gynecology residents reported an increase in didactics, while internal medicine residents on average perceived a decrease of 3.4 hours per week. Interestingly, despite the introduction and implementation of video conferencing, which dramatically facilitated meetings across our network, perceived attending involvement decreased overall by 1.2 hours per week (Supplemental Table 3). With video conferencing, presenters can share screens from remote locations, peers can assemble in large groups in a safe manner, and meetings can be recorded for viewing outside of scheduled time for convenience. However, despite these technological advantages and the decreased clinical responsibilities of some attendings and residents, we did not observe an increase in didactics as we expected.

In addition to the decrease in hours of weekly didactics, 53.6% of residents felt that the quality of didactics during the pandemic decreased, while 23.8% felt quality improved (Table 4). We observed a divergence between specialties in which 100% of pathology residents and 55% of obstetrics and gynecology residents reported an increase in quality, while 83.7% of internal medicine residents reported a worsening in quality. Examining this stark difference more closely, pathology residents felt that in addition to an improvement in the structure of didactics, there was more time to read and prepare for conferences, resulting in greater resident participation and overall preparedness.

Nationally, some programs have attempted to adapt by utilizing virtual videoconferences, inviting expert speakers, and recording conferences for later viewing for residents who are unable to attend.^6^ The Council on Resident Education in Obstetrics and Gynecology (CREOG) and Surgical Council on Resident Education (SCORE) started offering weekly supplemental lectures focusing on landmark journal articles, surgical technique, and exam related topics. Similarly, the Association of University Radiologists released a lecture series titled, “Diagnostic Radiology Resident Core Curriculum,” in an attempt to promote and standardize resident education and ameliorate the impact of the pandemic on resident education. These live lectures are delivered by leading educators in the field from prestigious academic institutions and have the potential to standardize education and reach a very wide audience. Nonetheless, distance learning cannot replicate many aspects of traditional educational forums; lack of true face-to-face interactions can limit engagement and investment on behalf of both presenters and listeners.^11^ Moreover, it is difficult to build the kind of interpersonal connections that are critical for professional development. Although the quality and utility of these lectures has not been directly assessed by the survey in this study, several respondents heralded the superior quality of these national lectures and felt this directly led to an improvement in didactics as a result of the pandemic.

Several questions in the survey sought to examine how residents’ clinical responsibilities and subsequent time for independent study changed during the pandemic. Within the healthcare network, internal medicine residents spent the most hours carrying for COVID-19 patients, with an average of 43.3-53.3 hours per week, followed by general surgery with 28-38 hours per week (Supplemental Table 2). Almost all pathology, radiology, and obstetrics and gynecology resident reported carrying 0-10 hours per week. Specific distribution of care of COVID-19 patients by specialties is currently not available in the literature; however, our findings are consistent with recently published reports and trends.^7^ The heavier burden of care of COVID-19 patients on internal medicine residents is concordant with the finding that they perceived to be working 5.1 more hours per week (SD=9.3), while all other specialties reported working less during the pandemic.

The change in the amount of time residents reported studying was inversely correlated to the clinical workload. Internal medicine residents, who reported working more, also reported studying 2.8 hours less per week, while general surgery, obstetrics and gynecology, pathology, and radiology residents reported studying 1.6, 5.8, 2.7, and 3.7 more hours per week, respectively (Table 2). Interestingly, only a portion of the extra time away from clinical work was redirected to studying independently across all specialties.

In addition to examining the immediate effects of the pandemic on education, this study sought to determine whether residents had long-term concerns about effects on their training. A large proportion of residents expressed concern that the pandemic diminished their preparedness to become an attending, however, few felt that they would be unable to complete graduation requirements or would require an additional year of training (Table 5). These trends were observed regardless of resident specialty. This seemingly incongruent lack of concern about achieving graduating requirement may be due to increasing flexibility on the part of accreditation societies; for example, the American Board of Surgery decreased the number of required operative cases.^9^

There are several limitations to this study. In order to achieve a good response rate, we conducted this study within our Healthcare Network. A benefit of this was that as a healthcare network in one of the earliest areas affected by COVID-19, we are able to analyze its effects in a timely fashion that may benefit other geographic areas affected similarly in the future. However, the results of this study may not be generalizable to residents in other programs in the United States. While residents in states with lower incidence of COVID-19 may not be as significantly impacted as residents at our institution, continued evolution of the COVID pandemic and the rise of new epicenters of disease may make these results more generalizable over time. Another limitation is the presence of recall bias; residents were asked to recount hours and experiences prior to the pandemic and compare them to the present time. This type of design can result in both perception bias as well as historical bias for which we cannot account.

Despite its limitations, these results are integral, as they highlight the varied but significant impacts of the pandemic on resident education. Different specialties were affected in diverse ways, and just as before the pandemic, education should be tailored to each specialty. Moving forward, program directors and leaders within each field should be cognizant of the changing needs of the residents within their specialty, keeping in mind that COVID-19 affected the amount of time spent on clinical work and studying uniquely based on the specialty. As we work to adapt quickly to evolving and previously unforeseen circumstances, we can learn from the experiences of our peers and colleagues.

## Conclusions

This study sought to examine the effects of the first wave of the COVID pandemic on residents within a healthcare network in the Northeast United States. The majority of residents reported diminished quality of education, time spent on organized didactics, and attending physician involvement in academics as a result of the COVID pandemic. Thirty-two percent of residents were concerned that the pandemic decreased their preparedness to become an attending. 13% were worried about completing graduation requirements, and 3% felt they would need an additional year of training. Unsurprisingly, residents in varying specialties were affected in different ways, illustrating that it is critical to develop tailored curriculum changes and create innovative opportunities for improving education, while continuing to encourage attending involvement in resident education during this challenging time.

## Data Availability

All data is presented in tables.

